# Serum proteomic correlates of arts and cultural engagement: implications for novel biological pathways linking arts to health

**DOI:** 10.64898/2026.01.09.26343769

**Authors:** Pei Qin, Saoirse Finn, Daisy Fancourt

## Abstract

Arts and cultural engagement (ACEng) have been linked to numerous physical and mental health outcomes across the life course, but research into the underlying biological mechanisms remains embryonic. We characterized the serum proteomic signatures of ACEng using data from a general population sample of ∼6,000 adults (Mean age: 53, SD: 18) in the UK Household Longitudinal Study with proteome profiling. Cross-sectional associations between ACEng and 184 cardiometabolic and neurological proteins were explored using multivariable linear regression, followed by functional enrichment analysis. Mediation analyses assessed whether identified proteins explained associations between ACEng and 15 cardiovascular, respiratory, musculoskeletal, metabolic, neurological and mental health conditions across a median of 20 years of follow up. Geater ACEng was associated with lower levels of 12 proteins and higher levels of six proteins after adjusting for sociodemographic confounders and multiple test corrections. Enriched biological pathways included cell proliferation, receptor-mediated signalling, immune regulation and inflammatory signalling. ACEng was associated with reduced prospective risk of most tested health conditions, and 70% of proteins were associated with at least one health condition. Proteins mediated the association between ACEng and disease risk; for example, TN-R mediated 10.23% of the association with asthma, SKR3 mediated 10.39% of the association with COPD, while MSR1 showed the largest mediation effects for stroke (11.81%), hypertension (11.27%), and diabetes (12.73%). Overall, all significant proteins mediated 15.95–37.70% of the associations between ACEng and the tested health outcomes. Main findings were replicated in extensive sensitivity analyses. These findings present, for the first time, proteomic signatures of ACEng and deepen our understanding of the mechanisms by which ACEng is linked to human health and disease.

## Introduction

Arts and cultural engagement (ACEng), including both actively participating in arts activities and attending cultural performances, events and heritage sites, is increasingly being recognised as an important health behaviour that can support mental, physical, and cognitive health across the life course ^1,2^. Meta-analyses and longitudinal studies have demonstrated that ACEng is associated with better cardiovascular, neurological, and mental health outcomes ^3–8^, as well as a reduced risk of all-cause mortality ^9,10^. The mechanisms that link ACEng to health outcomes are proposed to operate across psychological, social, behavioural, and biological processes ^12^.

Psychologically, ACEng can support emotion regulation, lower stress, and foster positive emotions ^13–16^, as well as provide a sense of meaning and purpose in life, enhance life satisfaction, and promote flourishing ^4,14,17,18^. Socially, ACEng can reduce loneliness and social isolation and facilitate feelings of social bonding and community ^19–24^. Behaviourally, ACEng can help individuals develop self-efficacy and a sense of mastery in their lives, affecting other aspects of their lifestyle and behaviours ^18,25,26^.

One emerging line of research is considering the role of ACEng in modulating inflammation. For example, participating in arousing arts activities such as singing and drumming for short periods of time has been shown to increase levels of pro-inflammatory cytokines, including interleukin-6 (IL-6) and tumor necrosis factor alpha (TNF-α) as well as other inflammatory proteins such as the chemokine monocyte chemoattractant protein-1 (MCP-1), indicating general activation of immune system activity ^27–29^. These findings are supported by studies testing leukocyte activity, which have also shown temporary increases from short-term arousing arts experiences, as well as faster wound healing ^29–32^. Conversely, pilot studies of relaxing arts experiences such as visiting galleries have reported reduced inflammatory markers ^33^. However, over time (i.e. weeks or months of ACEng), pilot studies of repeated arts engagement (both arousing and relaxing) has been associated with reductions in inflammatory markers and shifts towards anti-inflammatory immune profiles ^27,34^. Deepening the link between ACEng and protein pathways, the first studies of ACEng in relation to human gene expression have identified effects of short-term arts experiences on protein-coding mRNAs (messenger RNA) ^35–40^. The challenge is that most of the studies to date have relied on small sample sizes, focused on specific arts activities (without considering the broader context of arts activities individuals may be exposed to in their lives), and tested only individual or small panels of proteins. The first molecular epidemiological studies of ACEng have provided some corroboration of these pilot findings. Cross-sectional associations of two national cohort studies found that individuals more engaged in either dance specifically or arts and cultural activities more broadly had lower trait levels of inflammatory markers such as fibrinogen, CRP and white blood cells ^41,42^. However, these studies have similarly relied on only a few specific proteins based on pre-existing hypotheses and available blood biomarker data, limiting the opportunity to discover novel protein pathways related to ACEng that could have implications for mental and physical health.

Recently, the integration of high-throughput molecular biomarkers into national cohort studies has provided a novel opportunity to test the relationship between ACEng and proteins at a much larger scale, addressing this research gap. Proteomics involves the simultaneous assessment of dozens or hundreds of proteins, enables much deeper characterisation of the biological signature of behaviours and discovery of novel mechanistic pathways. Social proteomics in particular is an emerging field, which seeks to elucidate how social and lifestyle factors in people’s lives, such as social isolation, loneliness, and leisure-time physical activity, are associated with the human proteome ^43–45^. But to our knowledge, no studies to date have explored the proteomic signatures of ACEng in humans or the role of proteins in the relationship between ACEng and chronic disease explicitly.

Therefore, in this study, we examined the associations between ACEng and 184 proteins across cardiometabolic and neurological panels in a nationally representative cohort of UK adults, and performed functional enrichment analysis to characterise their biological function. We further investigated the relationships between ACEng and 14 health conditions using longitudinal follow-up data, and explored whether these proteins may mediate the associations between ACEng and the health conditions. This work stands to make a unique contribution to the literature as it has the potential to reveal novel biological pathways through which ACEng influences mental, physical, and cognitive health.

## Methods Dataset

This study used data from Understanding Society, the UK Household Longitudinal Study (UKHLS), which is a panel survey of members of approximately 40,000 private households in the UK ^46^. It was launched in 2009, with participants being followed up annually collecting data on a variety of sociodemographic and health information, with rich information on ACEng.

Between 2010 and 2012, approximately 20,000 adult participants (aged 16 and over) received a nurse health assessment visit from a registered nurse approximately five months after their Wave 2 interview (and, in the case of British Household Panel Survey (BHPS) members, after their Wave 3 interview). A range of anthropometric, physical function, and biological measures were collected. Non-fasting blood samples (n=13,130) were mailed to the processing laboratory, where they were separated into different fractions and frozen within a few days. The full details of nurse visits, including eligibility criteria, sampling processes, and assaying is published elsewhere ^47^.

Of the 54,568 participants in wave 2 of the UKHLS, we excluded those without the assay results of proteins (n=48,388), leaving a sample of 6,180 who had proteomics data. Individuals with missing protein measurements across the entire panels (cardiometabolic and neurology proteins) were excluded, leaving a sample size of 6,169 in the cardiometabolic dataset and 6,077 in the neurology dataset. ACEng exposures and covariates were obtained from the wave 2 adult survey. After further excluding participants with missing data on ACEng (n=103), the final analytical sample included 6,066 in the analysis of ACEng and cardiometabolic proteins and 5,974 for the neurology proteins (Figure 1).

**Figure 1.**
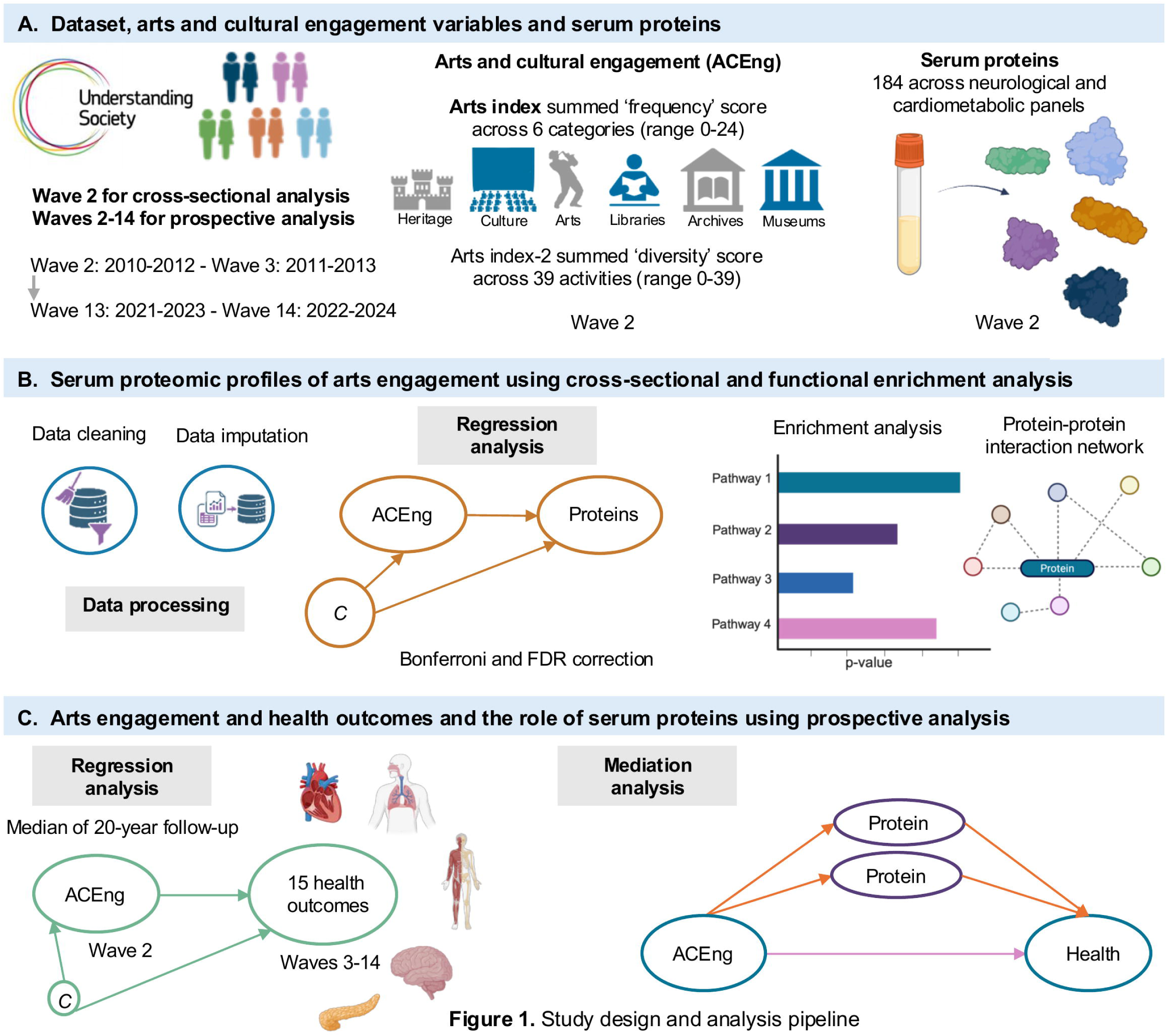
Methods and analysis pipeline. **(a) Dataset, arts and cultural engagement variables and serum proteins.** We used data from Understanding Society (also known as the UK Longitudinal Household Survey) for approximately 6,000 people. We used wave 2 for cross-sectional analyses for how arts and cultural engagement (ACEng) was associated with proteins and utilised longitudinal data to assess how ACEng was associated with health outcomes, and how proteins may mediate such associations. We created two indexes for ACEng capturing frequency and diversity of engagement and tested the associations with 184 serum proteins across neurological and cardiometabolic panels. **(b) Serum proteomic profiles of arts engagement using cross-sectional and function enrichment analysis.** After data cleaning and imputation, we ran linear regression analyses to assess how ACEng was associated with the 183 proteins, accounting for multiple testing using FDR and Bonferroni. We then performed functional enrichment analyses on significant proteins and assessed the protein-protein interaction network. **(c) Arts engagement and health outcomes and the role of serum proteins using prospective analysis.** Using a median follow-up of 20 years, we assessed how ACEng was associated with fourteen health outcomes, we then performed single and multiple mediation analyses to assess which ACEng-related proteins mediates the associations between ACEng and health outcomes.

Ethical approval for UKHLS was obtained from the University of Essex Ethics Committee and the National Research Ethics Service, and all participants provided informed consent. Figure 1 outlines the methods and analysis pipeline.

## Proteomics measurement

In 2020, a subset of serum samples (n=6,180) was sent to Olink® Proteomics (Uppsala, Sweden) for proteomic analysis ^48^. A total of 184 proteins were measured from two panels, with 92 from the cardiometabolic panel and 92 from the neurology panel. The proteins were measured using the Proximity Extension Assay (PEA) technology. A target protein is detected by its binding to two oligonucleotide-linked antibodies which have a DNA molecule artificially attached. The binding of the antibodies to the protein of interest brings close proximity for quantitative polymerase chain reaction (qPCR) which quantifies the level of the protein and yields a Ct value that indicates the amount of protein. The protein measurements are adjusted for batch effects and converted to NPX values (Normalised Protein eXpression) to quantify protein abundance. NPX values are based on log2-transformed Ct values, whereby a 1-unit increase corresponds to a doubling of the protein level.

## Arts and cultural engagement

Respondents were asked how often they had engaged in arts participation, cultural attendance, and visits to libraries, archives, museums, and heritage sites during the previous 12 months. Arts participation was assessed using a list of 14 activities, including dancing, singing, playing a musical instrument, reading for pleasure, photography, digital arts, painting, and crafting. Cultural attendance included 14 types of events, such as art exhibitions, street art displays, musical concerts, opera, and ballet. Heritage visits comprised eight types of historical sites, including historical parks or gardens and monuments such as castles. For library, archive, and museum visits, individuals were asked whether they had visited any of these places at least once during the last 12 months. A full list of the ACEng activities is presented in Table S1.

Creating an overall index of arts participation is complex, as individual behaviours can be extremely heterogenous in terms of frequency and diversity of engagement. While everyday ACEng can include different art forms (e.g., music, crafts, dance), modes of delivery (e.g., online, offline, self-led, led by an artist), and engagement types (e.g., actively doing, passively watching), they all share core common underlying active ingredients including multi-sensory stimulation, cognitive stimulation, imagination and aesthetic engagement ^11^. Therefore, even though there is variety in the types of activities people undertake, considering ACEng behaviours as a collective has strong precedence in health behaviour research (as is also common for other health behaviours such as physical activity). To ensure that decisions regarding the construction of our ACEng index did not overly bias results, we constructed two different indices and considered the stability of findings across the two. Both indices considered frequency and diversity of engagement together to give a more comprehensive overview of people’s ACEng behaviours, the first prioritised frequency within the index and the second prioritised diversity.

Our first ACEng index (hereafter ‘arts index’) summed the frequency score across each of the six categories for which people were asked about their patterns of engagement (arts participation, cultural attendance, library, archives, museum, and heritage visits). Frequency of engagement in each activity was categorised into five groups: ‘0 = not engaged’, ‘1 = once/twice in the last 12 months’, ‘2 = less often than once a month but at least three or four times a year’, ‘3 = less often than once a week but at least once a month’, and ‘4 = at least once a week’. Scores ranged from 0-24, with higher scores indicating greater frequency across more activities (thereby combining frequency and diversity).

Our second ACEng index (‘arts index-2’) leveraged the extremely rich data on all 39 types of arts activities asked about in the dataset, summing up the number of overall activities people reported doing. This provided an overall assessment of diversity of engagement (range, 0-39), but one that inherently captured frequency too given that high diversity necessitates more frequent overall ACEng engagement.

## Assessment of long-term health conditions

To gain deeper insights into the impact of ACEng on health and to elucidate potentially causal pathways linking ACEng to disease, we used wave 3 to wave 14 (approximately 11 years of follow-up) to ascertain the incident health conditions and further explore the role of serum proteomics in the relationship between ACEng and health. In UKHLS, participants’ self-reported long-term conditions included a pre-specified list of 17 conditions (asthma, arthritis, congestive heart failure, coronary heart disease, angina, heart attack or myocardial infarction, stroke, emphysema, hyperthyroidism, hypothyroidism, chronic bronchitis, any kind of liver condition, cancer or malignancy, diabetes, epilepsy, high blood pressure, and clinical depression), with dementia added since wave 14, captured in response to the question “*Has a doctor or other health professional ever told you that you have any of these conditions?*”. Most of the outcomes were censored up to wave 14 but hyperthyroidism and clinical depression was up to wave 9.

In the present analysis, the outcomes of interest included asthma, Chronic Obstructive Pulmonary Disease (COPD), Cardiovascular Disease (CVD), Ischaemic Heart Disease (IHD), stroke, heart failure, arthritis, hyperthyroidism, liver conditions, cancer, hypertension, diabetes, epilepsy, clinical depression, and dementia. Participants who reported being diagnosed with heart failure, coronary heart disease, angina, or stroke were classified as having CVD. Participants who reported being diagnosed with coronary heart disease or angina were classified as having IHD. Having emphysema or chronic bronchitis were classified as having COPD. When performing the analysis on health conditions, we only included the participants at wave 2 that had at least one follow-up between wave 3 and wave 14.

For each health outcome, the time-to-event data for new cases were calculated based on the intervals between baseline (wave 2) and the first reported event in the follow-up questionnaires (wave 3 to wave 14). Since these chronic conditions could only be identified during the periodic follow-up waves, the event date was determined according to the specific survey wave in which the event was first reported. The follow-up duration of the disease was then calculated as the time until the first diagnosis of the disease occurring after study entry up to the latest wave with complete data on the health conditions variables. Dementia was measured in wave 14 only, so we only generated the cases of dementia at wave 14 but not the follow-up duration.

## Covariates

Directed acyclic graphs (DAGs) were used to identify the minimally sufficient adjustment set, while avoiding covariates that could lie on the causal pathway and thereby partially mediate associations between ACEng and protein abundance ^49^. Covariates included age (years) and its quadratic term, sex (male, female), ethnicity (ethnic minority, White ethnicity), marital status (single/never married, married/cohabitating, separated/divorced/widowed), education level (no qualification, other, GCSE or below, A-level, Degree or above), employment status (not employed, employed), gross household income (quintiles), and area deprivation (quintiles using the 2019 Index of Multiple Deprivation (IMD)). Previous research suggests there is a bidirectional relationship between ACEng and other health behaviours. So health behaviours were not included in the main analyses. We also did not consider depression and other health conditions as confounders, as they are well-evidenced to lie on the causal pathway.

## Statistical analysis

After imputing the missing protein data using the median value, due to less than 2% missing data, the protein levels were standardized to mean of 0 and a standard deviation (SD) of 1 for analysis. To assess the cross-sectional associations between ACEng with each protein, we used linear regressions in a minimally adjusted model which included age, age^2^, sex, ethnicity, marital status, education level, employment status, income, and area deprivation. To account for multiple testing, we applied the Bonferroni correction method to indicate statistical significance (p = 2.717 × 10^−4^ (.05/184)). Restricted cubic splines were used to explore the linear and nonlinear dose-response relationship between ACEng and significant proteins, with three knots at 25%, 50%, and 75%.

Cox proportional hazards models were used to identify the significant proteins associated with 14 health outcomes, with a logistic regression model used for dementia. Analyses adjusted for age, age^2^, sex, ethnicity, marital status, education, employment status, income, and area deprivation. We also performed Cox proportional hazards models to explore the associations between ACEng and health outcomes (logistic regression model for dementia) adjusted for the same confounders.

We examined the mediating role of proteins in the associations between ACEng and each health outcome. Proteins that showed significant associations with ACEng in linear regression (with Bonferroni correction) were considered potential mediators. Two models were estimated: a multivariable linear regression model for proteins, conditioned on ACEng as the exposure, and a multivariable Cox regression model for all health outcomes, except dementia, which was analysed using logistic regression. Both models adjusted for minimally sufficient confounders (Model 1). The 95% confidence intervals of our estimates were generated using the percentile bootstrapping inference method, with 1000 bootstraps in each procedure and a random seed for reproducibility purposes. Mediation analysis was conducted using the ‘CMAverse’ R package ^50^. Additionally, all the protein mediators with significant effects in the one protein simple mediation were included in a multiple mediators mediation model to calculate the combined mediating effect.

Functional enrichment analysis was performed on the proteins that were significantly associated with ACEng (‘arts index’). We explored diverse biological pathways and several databases using Gene Ontology (GO) Biological Process, Molecular Function, Kyoto Encyclopedia of Genes and Genomes (KEGG), Reactome (REAC), and WikiPathways (WP). This was done to assess the biological mechanisms that ACEng may be involved in regarding the development of health outcomes, namely the biological functions of the identified proteins. We adopted FDR (‘False Discovery Rate’) correction for enrichment analyses to adjust for multiple comparisons. The gprofiler2 package in R was used to perform enrichment analysis ^51^. The protein–protein interaction (PPI) network of the proteins of interest was constructed using STRING ^52^. The cytoHubba plugin ^53^ in Cytoscape was adopted to identify hub proteins by the Maximal Clique Centrality (MCC) method ^54^

In sensitivity analyses, we repeated all analyses but using the ACEng ‘diversity’ index (arts index-2). We also conducted further sensitivity analyses by excluding those aged less than 40 years due to research suggesting this age to be a period of substantial biological ageing ^55^.

The analyses were conducted using R, version 4.3.2 (R Foundation for Statistical Computing) and all testing was two-tailed.

## Results

### Participant characteristics

Across our two protein samples (n=6,066 participants for cardiometabolic; n=5,974 for neurology proteins), the average age was 53 years (SD=18), 60% were female and most participants were of white ethnicity (96%) (Table 1). Additionally, 34% had a degree or above, 58% were married/cohabitating, 18% were current smokers, and 62% reported drinking alcohol more than once or twice a week times per week in the last 12 months.

**Table 1.**
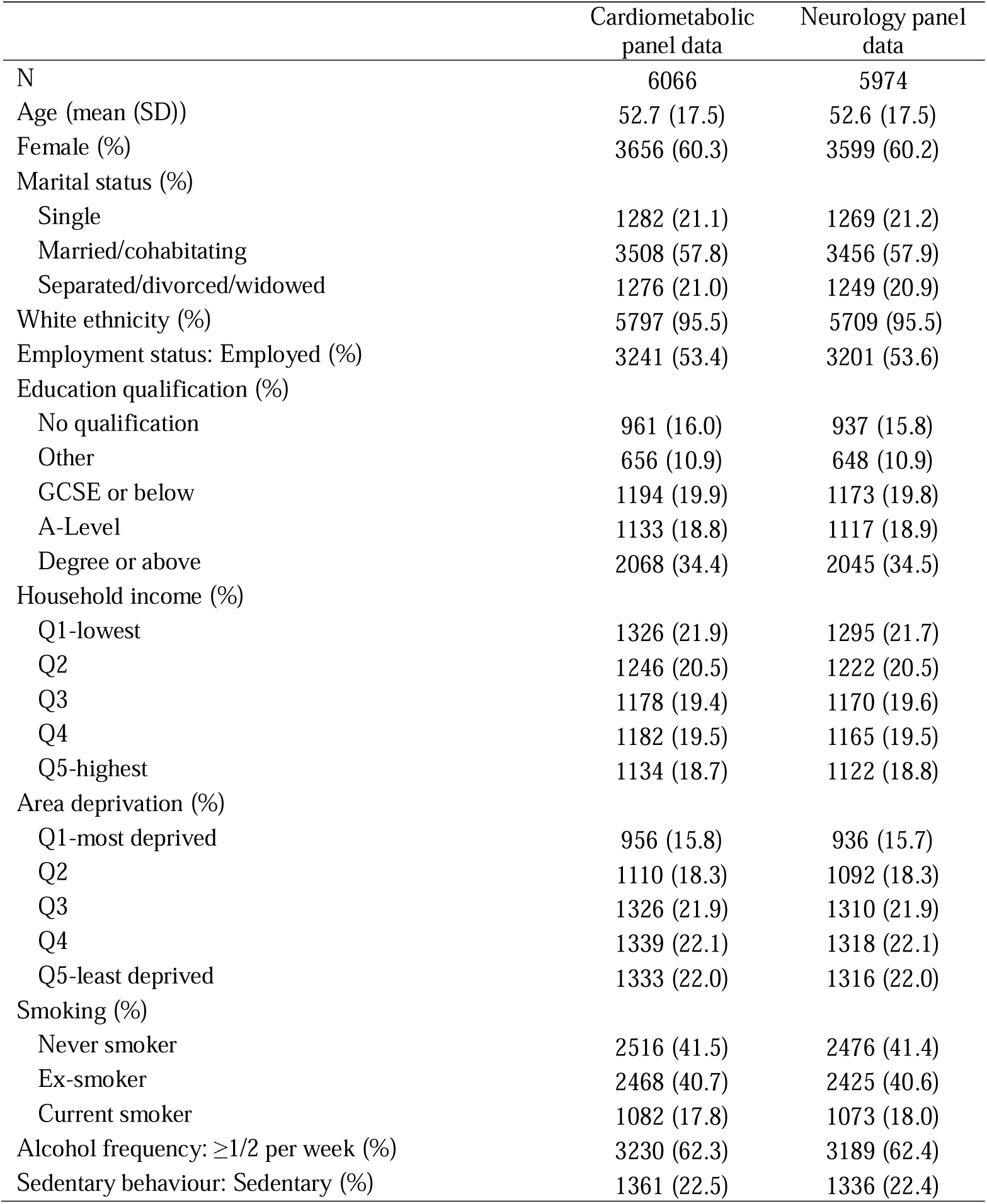
Participants characteristics

### Associations between arts and cultural engagement and serum proteomic markers

After adjusting for demographic and socioeconomic confounders (Model 1), greater ACEng (‘arts index’) was associated with lower levels of 12 serum proteins (SKR3, EFNA4, ICAM1, GFR-alpha-1, MSR1, LCN2, CST3, OSMR, TIMP1, REG1A, EDA2R, VWC2) and higher levels of six proteins (BCAN, NCAN, TN-R, IL7R, NTRK3, GDF-8) at the Bonferroni-corrected threshold (Figure 2a).

**Figure 2.**
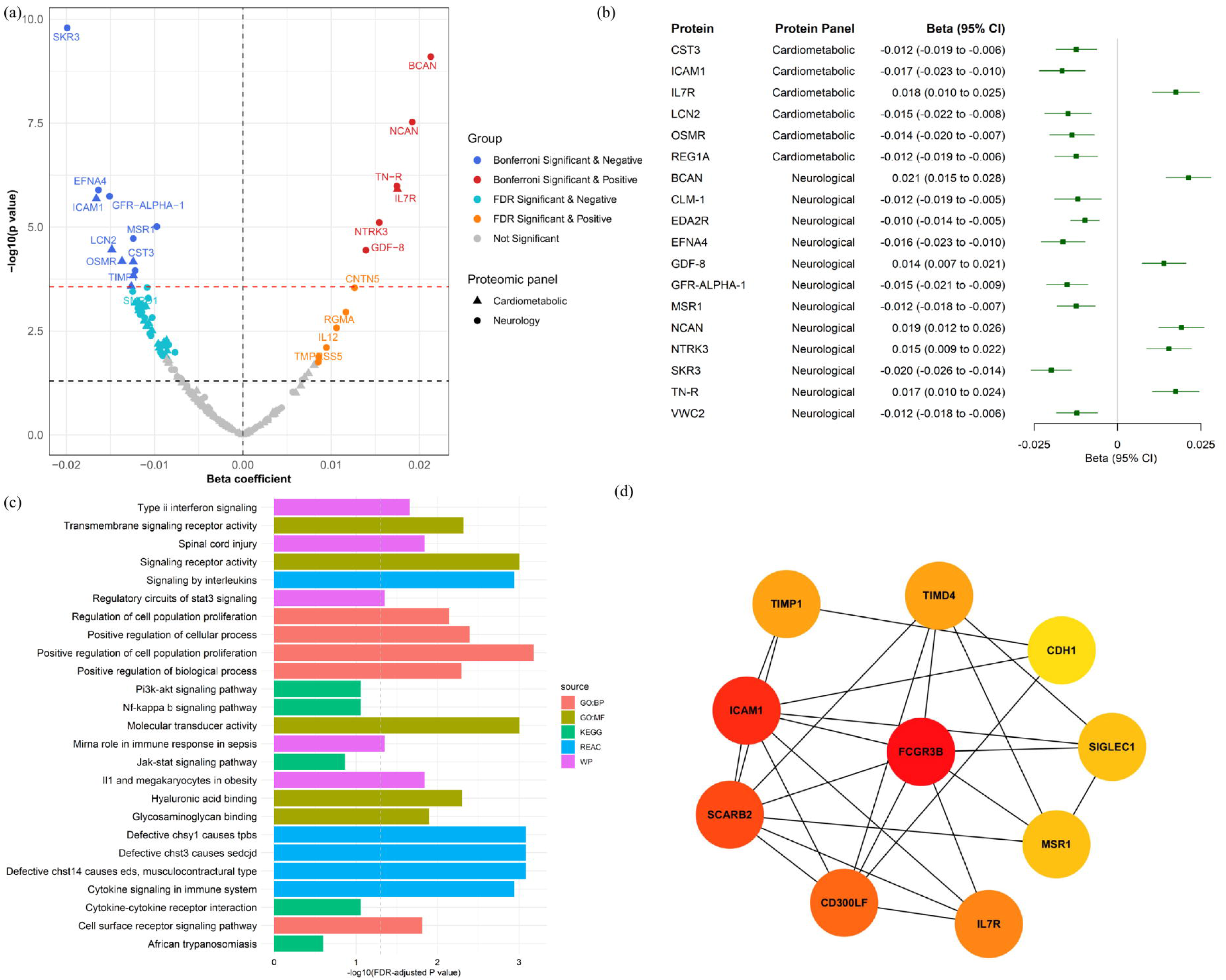
Serum proteomic profiles of arts and cultural engagement ‘frequency’ index (arts index). (a) **Volcano plots showing cardiometabolic and neurological proteins associated with arts index, highlighting strongly associated proteins.** The x-axis represents beta coefficients from the linear regressions, and the y-axis represents −log10(p-values). The red horizontal line shows the p-value for the Bonferroni correction, which is used to control for family-wise error rate (p<2.72×10^-4^ considered statistically significant). The black horizontal line shows the p-value at 0.05. The associations were all adjusted for age, age^2^, sex, ethnicity, marital status, education, income, employment status, and area deprivation. **(b)**Forest plot of the association between arts index and proteins. The proteins that were significant after Bonferroni correction. (c) Functional enrichment analyses of serum proteins associated with arts index. (d) Top 10 hub proteins using the CytoHubba MCC clustering algorithm.

BCAN demonstrated the strongest positive association with ACEng (β: 0.021, 95% CI: 0.015 to 0.028, p=7.91E-10), followed by NCAN (β: 0.019, 95% CI: 0.012 to 0.026, p=2.96E-08), IL7R (β: 0.018, 95% CI: 0.010 to 0.025, p=2.96E-08), and TN-R (β: 0.017, 95% CI: 0.010 to 0.024, p=2.96E-08) (Figure 2b).

SKR3 demonstrated the strongest negative association with ACEng (β: -0.020, 95% CI: -0.026 to - 0.014, p=1.61E-10), followed by ICAM1 (β: -0.017, 95% CI: -0.023 to -0.010, p=9.45E-05), EFNA4 (β: -0.016, 95% CI: -0.023 to -0.010; p=2.36E-05), and GFR-alpha-1 (β: -0.015, 95% CI: -0.0021 to - 0.009, p=2.78E-05) (Figure 2b).

As shown in the restricted cubic splines, most proteins showed a linear relationship with ACEng (Figure 3).

**Figure 3.**
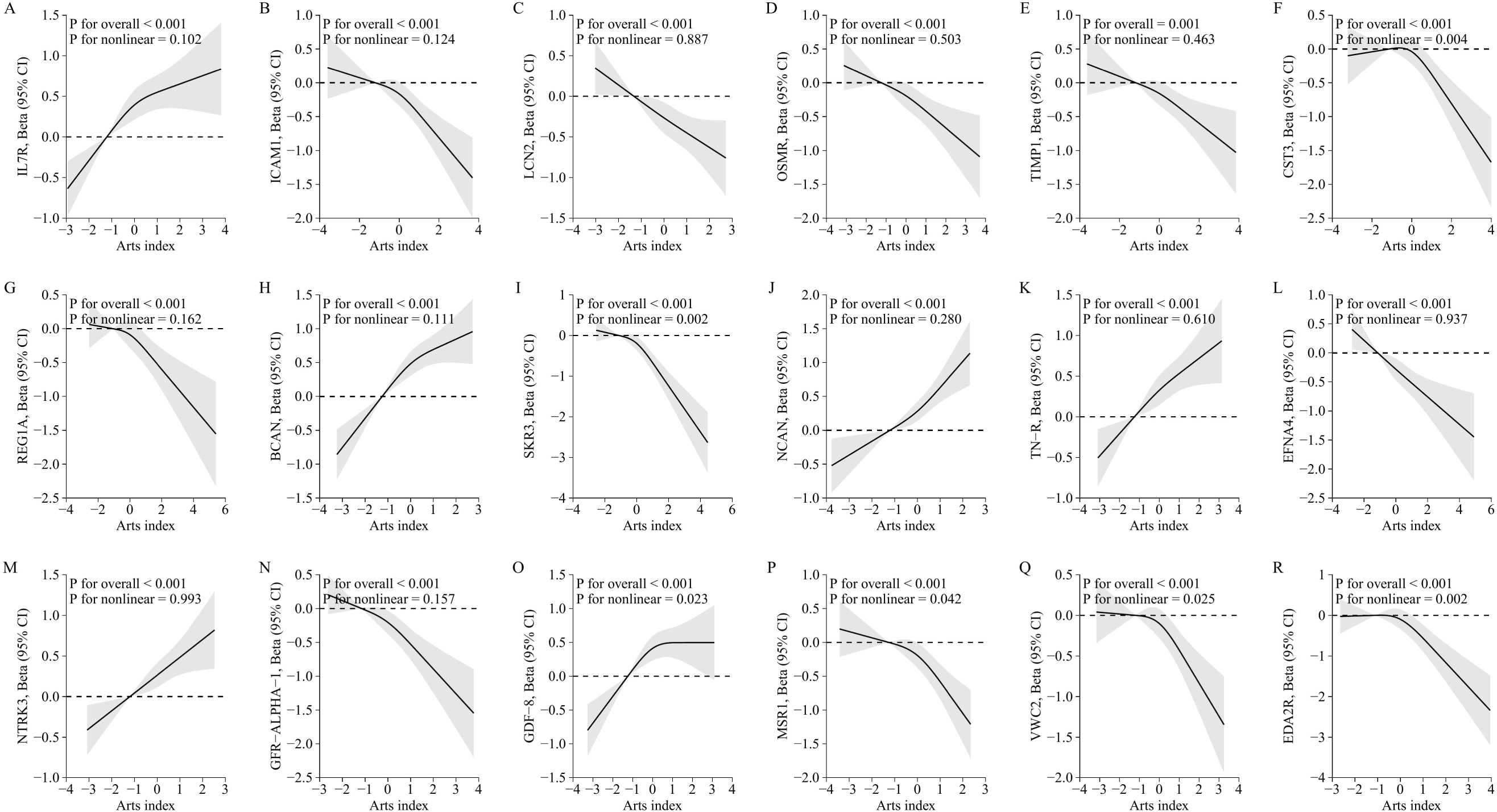
Dose-response relationship between arts and cultural engagement ‘frequency’ index (arts index) and serum proteins that were significant after Bonferroni correction.

### Pathway enrichment of serum proteomic signatures of arts and cultural engagement

Functional enrichment analysis was performed on the 18 proteins that were significantly associated with ACEng (‘arts index’) at the Bonferroni-corrected threshold (Figure 2c). Notably, the top enriched biological processes were mainly related to cell proliferation and receptor-mediated signalling, including positive regulation of cell population proliferation, positive regulation of cellular process, positive regulation of biological process, regulation of cell population proliferation, and cell surface receptor signalling pathway (Figure 2c; Table S2).

The REAC and WP enrichment highlighted pathways related to inflammatory signalling and immune regulation such as signalling by interleukins, cytokine signalling in the immune system, regulatory circuits of STAT3 signalling, miRNA roles in immune response in sepsis, type II interferon signaling, and IL1 and megakaryocytes in obesity (Figure 2c; Table S2).

Subsequently, we conducted a PPI analysis to elucidate the intricate connections among the 59 proteins related to ACEng that met FDR significance and identify key proteins within this network. Utilizing STRING, we generated a PPI network comprising 56 nodes and 75 edges (interactions) (Figure S2). To identify key hub proteins within the PPI network, the MCC algorithm in Cytoscape’s cytoHubba plugin was applied. FCGR3B, ICAM1, and SCARB2 exhibited the highest centrality, indicating the biological pathways related to immune and inflammatory processes, cellular stress response, cell adhesion, and extracellular matrix remodeling (Figure 2d).

### Associations between arts and cultural engagement and health outcomes

To explore the role of serum proteins in the association between ACEng (‘arts index’) and health outcomes, we first performed Cox proportional hazard regression models (except for dementia using a logistic regression model) association between arts index and 15 health outcomes. The sample size of all the outcomes ranged from 659 for dementia and 3,288 for hypertension. The median follow-up duration was 20 years. The incident cases ranged from 51 for hyperthyroidism to 1,306 for hypertension (Table S3).

After adjusting for sociodemographic covariates (Model 1), higher ACEng was associated with decreased risk of all health outcomes except for heart failure, liver disease, and epilepsy: asthma (HR: 0.963, 95% CI: 0.941 to 0.987), COPD (HR: 0.896, 95% CI: 0.858 to 0.935), CVD (HR: 0.957, 95% CI: 0.933 to 0.980), IHD (HR: 0.943, 95% CI: 0.916 to 0.970), stroke (HR: 0.949, 95% CI: 0.903 to 0.997), arthritis (HR: 0.955, 95% CI: 0.939 to 0.971), hyperthyroidism (HR: 0.912, 95% CI: 0.840 to 0.990), cancer (HR: 0.972, 95% CI: 0.948 to 0.996), hypertension (HR: 0.961, 95% CI: 0.946 to 0.976), diabetes (HR: 0.931, 95% CI: 0.905 to 0.957), clinical depression (HR: 0.951, 95% CI: 0.927 to 0.975), and dementia (HR: 0.813, 95% CI: 0.667 to 0.992) (Figure 4a).

**Figure 4.**
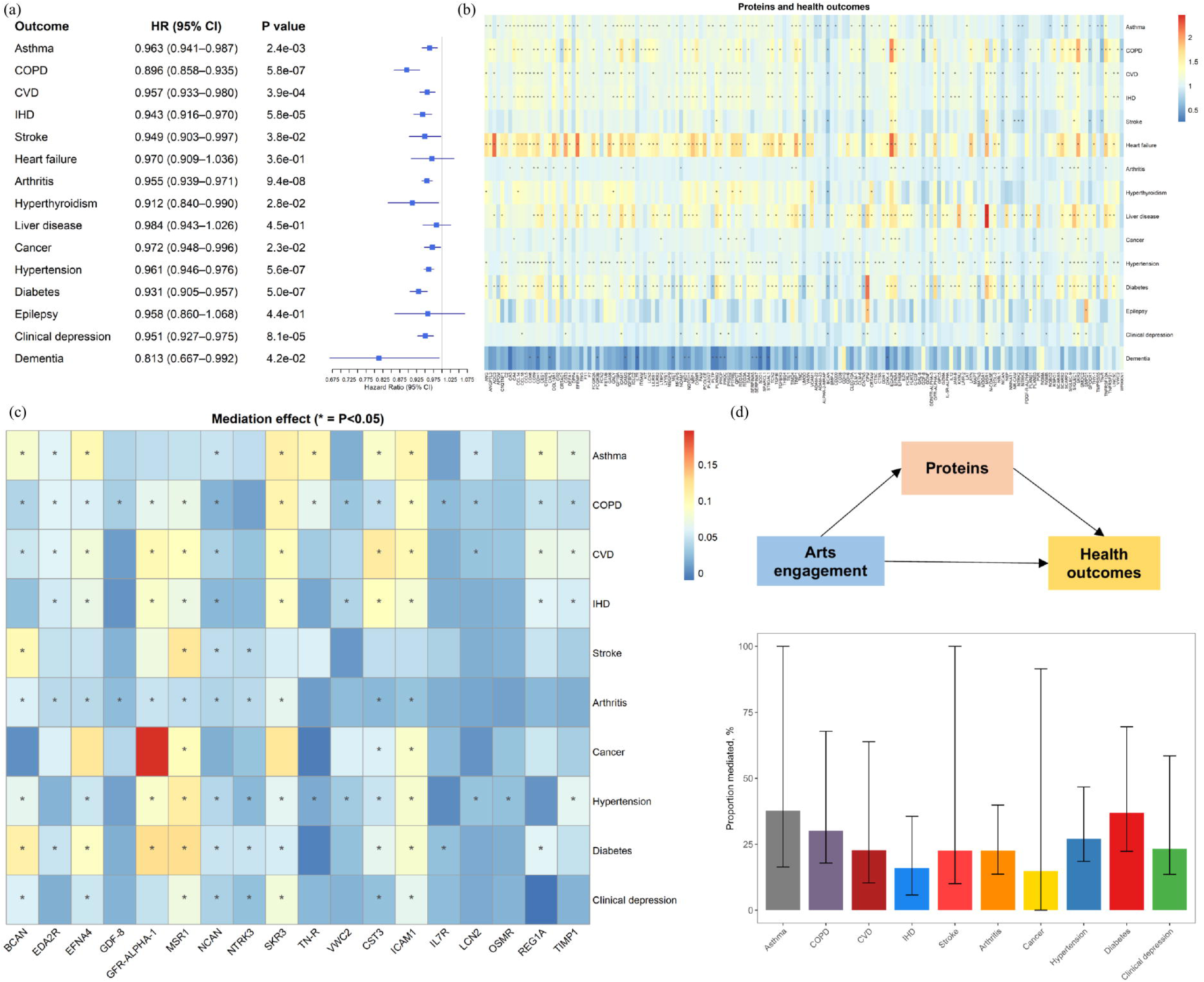
Arts and cultural engagement ‘frequency’ index and health outcomes and the role of serum proteins. (a) **Forest plots showing arts index and associations with 14 long-term health outcomes.** Cox regression models were conducted to generate the Hazard ratios and 95% CIs of arts index and health outcomes (except for dementia). The models were all adjusted for age, age^2^, sex, ethnicity, marital status, education, income, employment status, and area deprivation. (b) **Heatmap showing the associations between serum proteins and health outcomes.** The models were all adjusted for age, age^2^, sex, ethnicity, marital status, education, income, employment status, and area deprivation. (c) **Single mediation analyses of serum proteins in the associations between arts index and health outcomes.** The heatmap only shows the percentage of mediation for the health outcomes that had a significant indirect effect of the protein in the associations between arts index and health outcomes. Those proteins that were non-significant across all outcomes are not shown. The value is the percentage of mediation of each protein and those which were significant in the mediation analysis are marked as “*”. The mediation models were all adjusted for age, age^2^, sex, ethnicity, marital status, education, income, employment status, and area deprivation. (d) **Multiple mediation analyses of all identified significant proteins in the associations between arts index and health outcomes.** The models were all adjusted for age, age^2^, sex, ethnicity, marital status, education, income, employment status, and area deprivation.

### Associations between serum proteins and health outcomes

Of the 184 proteins explored, a series of proteins were significantly associated with increased risk of health outcomes after adjustment of sociodemographic covariates (Figure 4b; Table S4). For example, 66 proteins were significantly associated with risk of incident asthma, 47 were associated with COPD, 47 were associated with CVD, 45 with liver disease, 146 with diabetes, 63 with hypertension, and 19 with clinical depression (FDR corrected p-value< 0.05).

### Mediation analysis of proteins in the associations between arts and cultural engagement and health outcomes

We explored the significant mediation effect of each of the 18 proteins significantly related to ACEng and 15 health outcomes. Among these proteins, SKR3 contributed to a large proportion of mediation for asthma (10.92%), COPD (10.39%), CVD (9.72%), IHD (8.96%) and depression (7.81%). MSR1 had the highest proportion of mediation effect in stroke (11.81%), hypertension (11.27%), and diabetes (12.73%). GFR-alpha-1 contributed to the large proportion of mediation for CVD (10.01%), IHD (8.70%), hypertension (8.71%), and diabetes (12.67%). ICAM1 had a high proportion of mediation effect in asthma (10.21%), CVD (10.15%), cancer (8.65%), COPD (8.92%), diabetes (8.42%), and hypertension (8.45%). CST3 contributed the largest proportion of mediation effect for CVD (11.48%) and IHD (9.48%) and TN-R contributed the largest proportion for asthma (10.23%). Additionally, BCAN mediated 10.86% and 10.01% of the association for diabetes and stroke (Figure 4c; Table S5).

Collectively, these proteins partially mediated 37.70% (16.38%-217.44%) of asthma risk, 30.17% (17.87%-67.84%) of COPD risk, 22.75% (10.34%-63.89%) of CVD risk, 15.95% (5.76%-35.60%) of IHD risk, 22.57% (10.05%-122.21%) of stroke risk, 22.56% (13.66%-39.86%) of arthritis risk, 27.07% (18.51%-46.71%) of hypertension risk, 36.92% (22.33%-69.53%) of diabetes risk, and 23.25% (13.57%-58.51%) of clinical depression risk related to ACEng (Figure 4d; Table S6).

## Sensitivity analysis

Sensitivity analyses using the second arts index (‘arts index-2’) were performed. The analysis yielded results consistent with the main analysis. After adjusting for sociodemographic covariates, 22 proteins were associated with arts index-2 after applying Bonferroni correction (Figure S3a and Figure S3b). BCAN, IL7R, GDF-8, and NCAN remained among the top 4 proteins that showed positive associations. SKR3, GFR-alpha-1, EFNA4, and ICAM1 remained the top 4 proteins that showed negative associations. Similar biological pathways related to inflammation and immune regulation were also identified by the arts index-2 enrichment analysis, such as cytokine signalling in the immune system and regulatory circuits of STAT3 signaling (Figure S3c; Table S7). The top protein hubs based on the PPI network also included ICAM1 and SCARB2 (Figure S2; Figure S3d). Linear relationships were also identified between arts index-2 and most of the significant serum proteins (Figure S4).

Except for the different results for cancer, arts index-2 showed consistently significant associations with lower risk of asthma, COPD, CVD, IHD, stroke, arthritis, hyperthyroidism, hypertension, diabetes, clinical depression, and dementia (Figure S5a). Significant mediation effects of certain proteins were also identified in ACEng and asthma, COPD, CVD, IHD, stroke, arthritis, hyperthyroidism, hypertension, diabetes, and clinical depression (Figure S5b; Table S8). Consistent findings were shown for the multiple protein mediation analyses (Figure S5c; Table S9).

We also performed sensitivity analysis by excluding those aged less than 40 years, with the sample size of 4,494 for cardiometabolic proteins panel and 4,565 for neurology proteins panel analysis. The results showed 21 proteins associated with arts index (P value <2.717 × 10^−4^), which included those proteins that were identified as significant in the main analysis (Figure S6).

## Discussion

Using a large population-based study with a comprehensive measurement of arts and cultural engagement (ACEng), our data-driven approach provided novel evidence of serum protein signatures of ACEng. In our main analyses, 18 proteins were cross-sectionally associated with greater ACEng in fully-adjusted models that held after Bonferroni correction. The top enriched biological pathways included cell proliferation, receptor-mediated signalling, immune regulation and inflammatory signalling. Greater ACEng was associated with a reduced future risk of asthma, COPD, CVD, IHD, stroke, arthritis, hyperthyroidism, cancer, hypertension, diabetes, clinical depression and dementia over a median of 20 year follow-up. Approximately 70% of proteins were significantly associated with at least one health condition. The identified proteins related to ACEng collectively mediated between 15.95–37.70% of the association between ACEng and health conditions. Additionally, our findings were also replicated in extensive sensitivity analyses. To our knowledge, this is the first study to explore the associations between ACEng and human protein signatures, where we have identified proteins and biological pathways that have not previously been linked to ACEng, or how ACEng impacts health.

Of the 17 shared proteins between the two ACEng indexes, ∼71% were part of the neurological panel. We found that greater ACEng was associated with higher NCAN and BCAN levels which are both primarily expressed in the brain. NCAN is implicated in early neural development, but its dysregulation has been linked to neurodegenerative processes seen in Alzheimer’s disease, such as altering of perineuronal nets, as well as to neuropsychiatric and neurological disorders including bipolar disorder, schizophrenia, depression and epilepsy ^56–58^. Indeed, higher NCAN levels have been correlated with better brain health, including better white matter integrity, greater whole brain and grey matter volume, as well as greater Aβ1-42 (amyloid-beta 1-42) levels, which may suggest lower deposits of amyloid-beta plaques in the brain ^59–61^. BCAN is believed to be involved in central nervous system processes in the mature brain, such as synaptic organisation and plasticity. Indeed, greater BCAN levels have been linked to greater grey and white matter brain volume, better cognitive function, and lower risk of dementia and depression, with individuals with cognitive impairment and Alzheimer’s disease showing lower levels of BCAN ^62–65^. This has highlighted how BCAN could be a candidate biomarker for brain ageing ^64^. In medical and neuroimaging studies, arts engagement (in conjunction with other social or leisure factors) has been linked to higher levels of Aβ1-42 measured in cerebrospinal fluids and lower levels of white matter damage in older adults ^66,67^. Therefore, our findings support and extend our understanding of the impact of ACEng on brain health and biology.

We also found evidence of ACEng affecting inflammatory processes and immune system regulation. For example, ICAM-1 is involved in immune and inflammatory responses, it is known for modulating leukocyte activity during acute infections and in chronic inflammation, and has been linked to conditions such as rheumatoid arthritis, cardiovascular diseases, cancers, asthma, as well as responses to respiratory infections, such as COVID-19 ^68,69^. Therefore, it could be proposed that lower ICAM-1 levels found in our analyses indicate that greater ACEng is associated with lower inflammation. This would corroborate the initial experimental and epidemiological evidence reviewed above, but provide a key extension by assessing large-scale proteomic panels and finding associations with novel inflammatory proteins.

In addition, we found support for ACEng impacting proteins implicated in vascular and metabolic regulation. Greater ACEng was associated with lower SKR3 (also known as activin-like kinase receptor 1 (ALK1)), which is believed to be involved in vascular development and angiogenesis ^70^, as well as allowing lipids into blood vessels ^71^. Indeed, ALK1 has been implicated in atherosclerosis ^72^, cerebrovascular diseases ^73^, and diabetic neuropathy ^74^. Our findings align with the small number of studies that have assessed how ACEng is associated with proteins involved in cardiometabolic processes, such as HbA1c ^42^, but we have identified novel proteins for how ACEng may impact inflammatory, vascular and metabolic functions.

Additional pathways found to be enriched included hyaluronic acid binding, cell surface receptor signaling pathway, regulatory circuits of STAT3 signaling, cytokine signaling in immune system, Type II interferon signaling, transmembrane signaling receptor activity, and IL-1 and megakaryocytes in obesity. Furthermore, across the top hub proteins identified in PPI analyses, across both indexes, ICAM1, SCARB2, TIMP1 and IL7R showed high centrality, indicating biological pathways related to immune and inflammatory processes, cell adhesion, lipid transport, and extracellular matrix remodeling. This suggests additional plausible biological mechanisms through which ACEng may be linked to health.

Positioning our findings in relation to broader emerging social proteomics work, recent studies have suggested social isolation is associated with higher plasma SKR3, and greater social support with lower plasma SKR3 and EFNA4, and higher TN-R ^44,45^. This mirrors our findings whereby we found lower serum SKR3 and EFNA4 from greater ACEng, and higher TN-R. This suggests that ACEng has a similar protein signature to favourable social connection profiles, indicating that leisure and social engagement may have similar biological processes. On the one hand, this is not surprising given that ACEng frequently involves social interaction. However, interestingly, despite ACEng containing active ingredients involving different intensities of physical exertion depending on the activity, proteins commonly associated with physical activity did not tend to overlap with our findings ^43^. This suggests that ACEng is a unique health-promoting leisure activity that is not purely driven by elements of physical activity. Our findings add to existing theoretical work, by exploring novel biological mechanisms linking ACEng to health ^12^. Indeed, the consistency of our findings across indices that prioritised frequency and diversity extends our understanding of the health impacts of ACEng in relation to social identity theory, which posits that identifying with more group memberships is beneficial to health ^75^.

The strengths of our present study included the use of a large UK adult cohort, encompassing a broad array of proteins, adopting a well-rounded measurement of arts engagement that involved a variety of activities and their frequency and generating a novel index to fully consider the frequency and diversity of ACEng, adjusting for a series of confounders, undertaking functional enrichment and PPI network analyses to explore biological pathways and testing consistency across a range of sensitivity analyses. However, it is important to note that the analyses of the relationship between ACEng and proteins were cross-sectional. Essentially, the findings suggest a protein signature of ACEng. This work is, nonetheless, an important step forwards, as it leverages high-throughput molecular biomarkers to identify candidate proteins that can now be prioritised in smaller-scale protein panels in experimental studies to extend the work presented here and test causality further. As such, our findings provide critical ground-work to generate novel causal hypotheses. Furthermore, we undertook the first longitudinal mediation analyses to test potential mediating effects of these proteins on long-term health outcomes that we identified as associated with ACEng. Utilising the longitudinal data from UKHLS with a median of 20 years’ follow up, we found that ACEng was associated with reduced risk of cardiovascular, respiratory, musculoskeletal, metabolic, neurological and mental health conditions. These findings corroborate multiple previous studies ^5,76–79^, as well as extending them to include several conditions not extensively studied in relation to ACEng such as asthma, arthritis and hyperthyroidism in population-level analyses. The ACEng-relevant proteins were further found to mediate the association between ACEng and most of these health outcomes, including COPD, CVD, IHD, stroke, hypertension, diabetes and clinical depression. These findings have important implications into the underpinning of the biological mechanisms that may link ACEng to human health and wellbeing and further support the rationale of conducting targeted intervention studies and implementing arts and cultural engagement promotion programs. Additionally, future interventions studies could adopt the novel biomarkers identified in our present study to monitor, understand, and predict the early effects of ACEng.

Our study has several additional limitations to consider. First, the data are not representative of ethnic minority groups or the entire UK, as Northern Ireland is not included. Second, we did not perform validation studies and could not make conclusions as to whether the findings can be validated in other datasets, as ACEng is extensively phenotyped in Understanding Society and not similarly measured by other potential cohorts. Future studies are essential to replicate our results. Third, the cross-sectional design to identify proteomic signatures of arts engagement was limited to determine causal relationships. We recommend that future epidemiological research on ACEng adopts advanced causal mediation approaches, examines temporality between ACEng and proteins, and assesses longitudinal patterns of ACEng and proteins. Forth, we cannot exclude the measurement error of ACEng and confounders from the self-reported data. Fifth, the proteomic measurement in the present study only included selected proteins in two panels of OLink platform. Future studies are recommended to explore proteome-wide relationships across larger protein panels and biological systems and leverage multi-omics strategies to uncover the biological mechanisms linking ACEng to health and disease. Finally, the health outcomes were ascertained by self-report in the follow-up waves and there was a relatively large proportion of missing data on self-reported health conditions. Future studies with health outcomes by linkage to the hospital records data are warranted to validate our findings.

## Conclusions

Overall, our study found novel serum proteins associated with ACEng in a large sample of adults living in the UK. The findings provide the first known data on the relationship between ACEng and the human proteome. We have demonstrated novel biological pathways associated with ACEng including cell proliferation, receptor-mediated signalling, immune regulation and inflammatory signalling, extending previous pilot research showing ACEng impacts biological processes relating to neurological, inflammatory and metabolic pathways. This helps to provide further biological plausibility and mechanistic evidence for the impact of ACEng on human health and opens opportunities for a new era of ‘leisure proteomics’ research.

## Data Availability

Data can be accessed via UKDS: https://datacatalogue.ukdataservice.ac.uk/series/series/2000053#abstract

## Acknowledgements

Funding for this research was provided as part of the Understanding Society Fellowship programme, a component of the Study’s Economic and Social Research Council award [ES/S007253/1]. This research was also supported by UK Research and Innovation [MR/Y01068X/1] and a Wellcome Trust Discovery Award [326117/Z/25/Z].

