## Supplemental (Figures) for "Serum proteomic correlates of arts and cultural engagement: implications for novel biological pathways linking arts to health"

**Supplemental files**

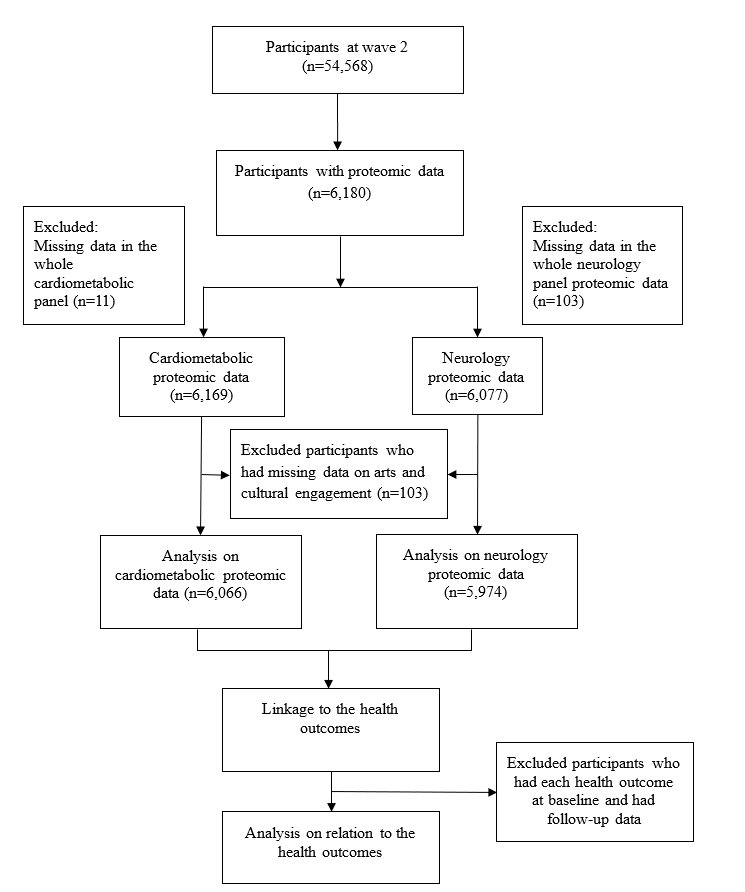

**Figure S1.** Participant selection.

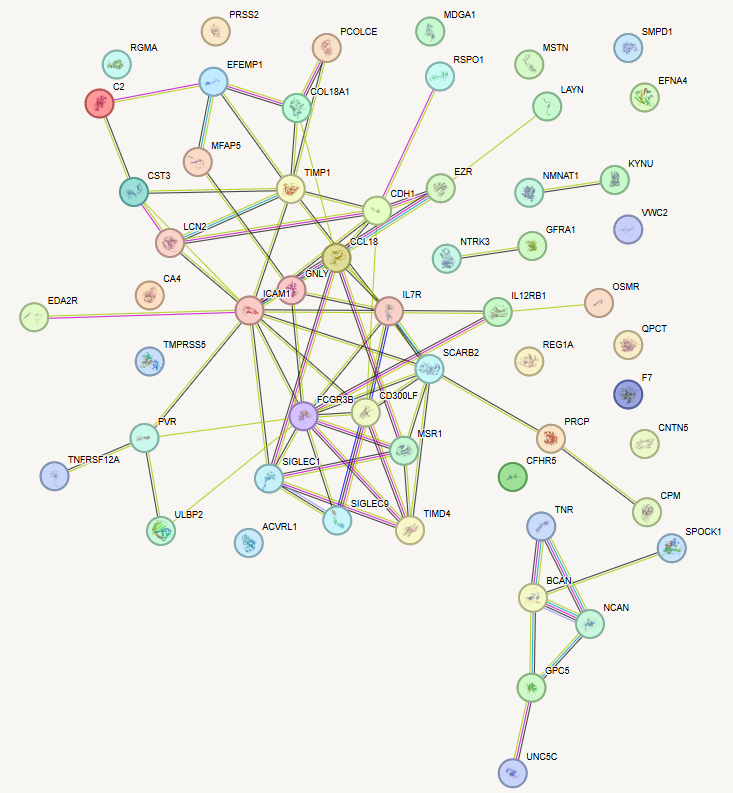

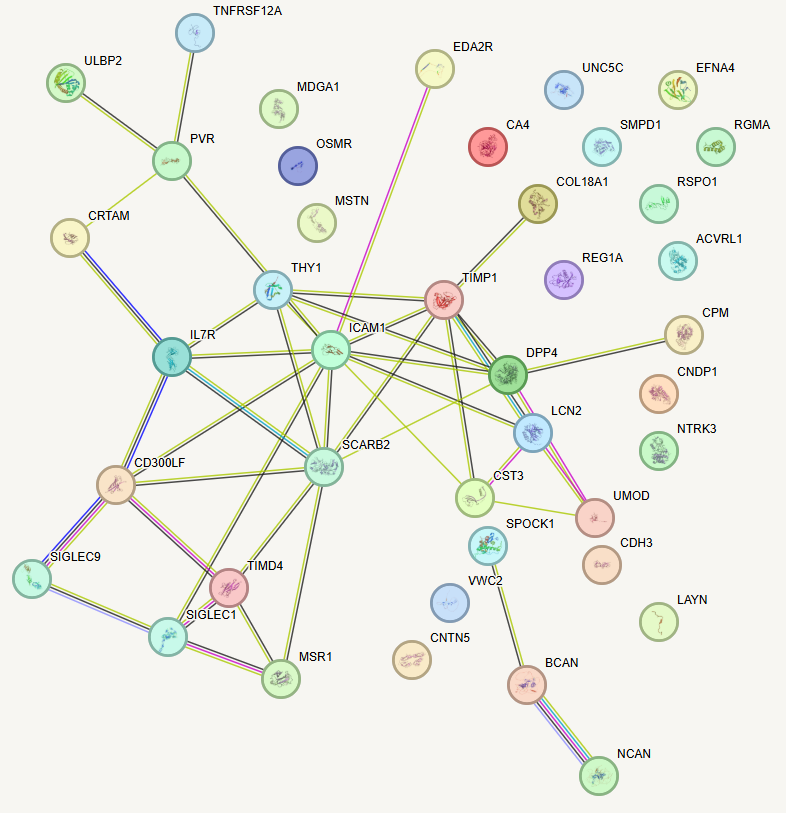

(b)

(a)

**Figure S2.** Protein–protein interaction (PPI) networks within the identified pool of 59 proteins related to arts index (a) and 47 proteins related to the arts index-2 (b) that met FDR significance. PPI network based on STRING.

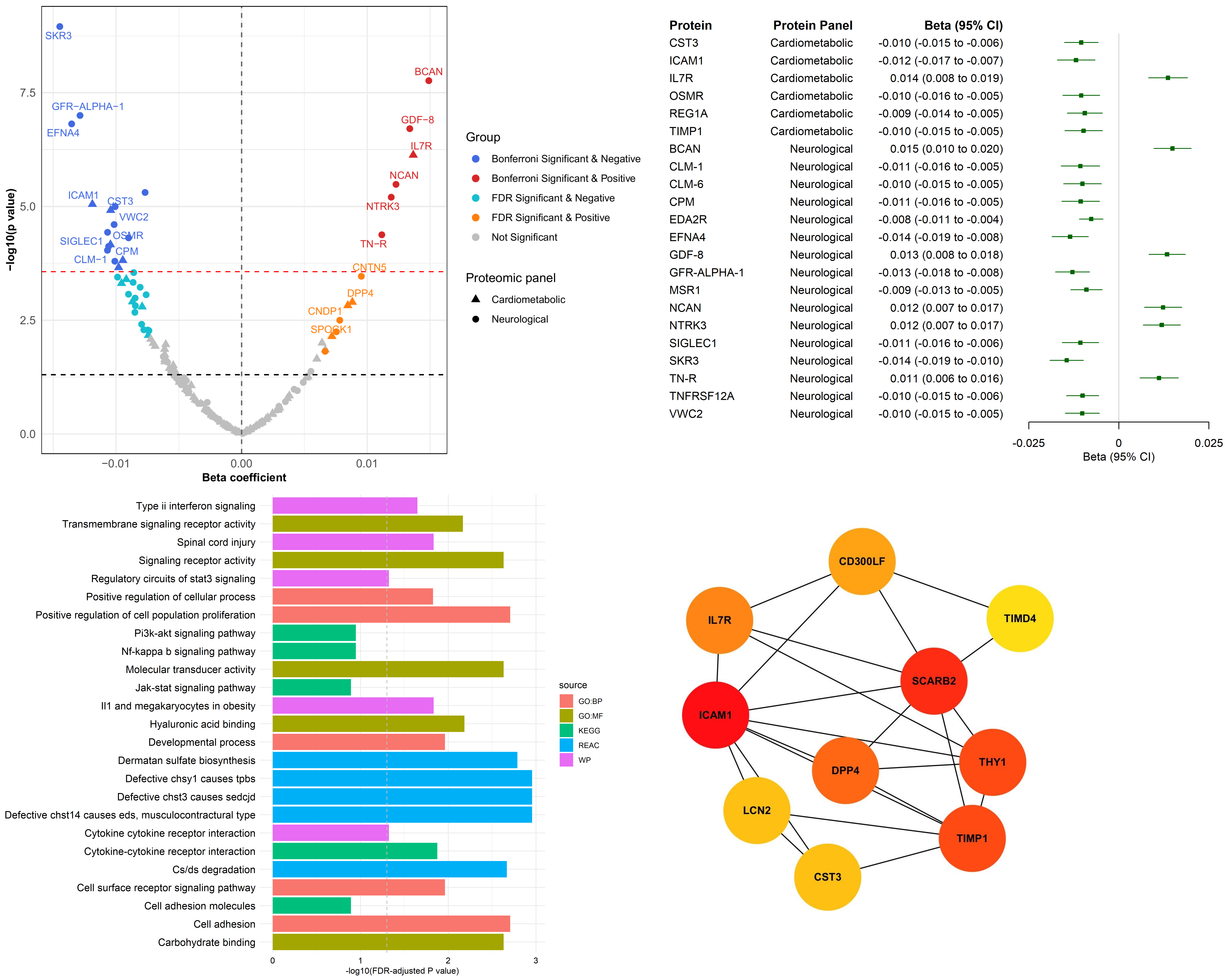

(b)

(a)

(c)

(d)

**Figure S3.** Serum proteomic profiles of arts index-2 as sensitivity analyses.
To test the robustness of our findings regarding our measurement approach for arts and cultural engagement, arts index-2 leveraged the extremely rich data on all 39 types of arts activities asked about in the dataset, summing the number of overall activities people reported doing. This provided an overall assessment of ‘diversity’ of engagement.

**(a) Volcano plots showing proteins associated with arts index-2, highlighting strongly associated proteins.** The x-axis represents beta coefficients from the linear regressions, and the y-axis represents −log10(p-values). The red horizontal line shows the p-value for the Bonferroni correction, which is used to control for family-wise error rate (p<2.72×10^-4^ considered statistically significant). The black horizontal line shows the p-value at 0.05. The associations were all adjusted for age, age^2^, sex, ethnicity, marital status, education, income, employment status, and area deprivation
**(b) Forest plot of the associations between arts index-2 and proteins.** The proteins that were significant after Bonferroni correction.
**(c) Functional enrichment analyses of proteins associated with arts index-2.**
**(d) Top 10 hub proteins associated with arts index-2 using the CytoHubba MCC clustering algorithm.**

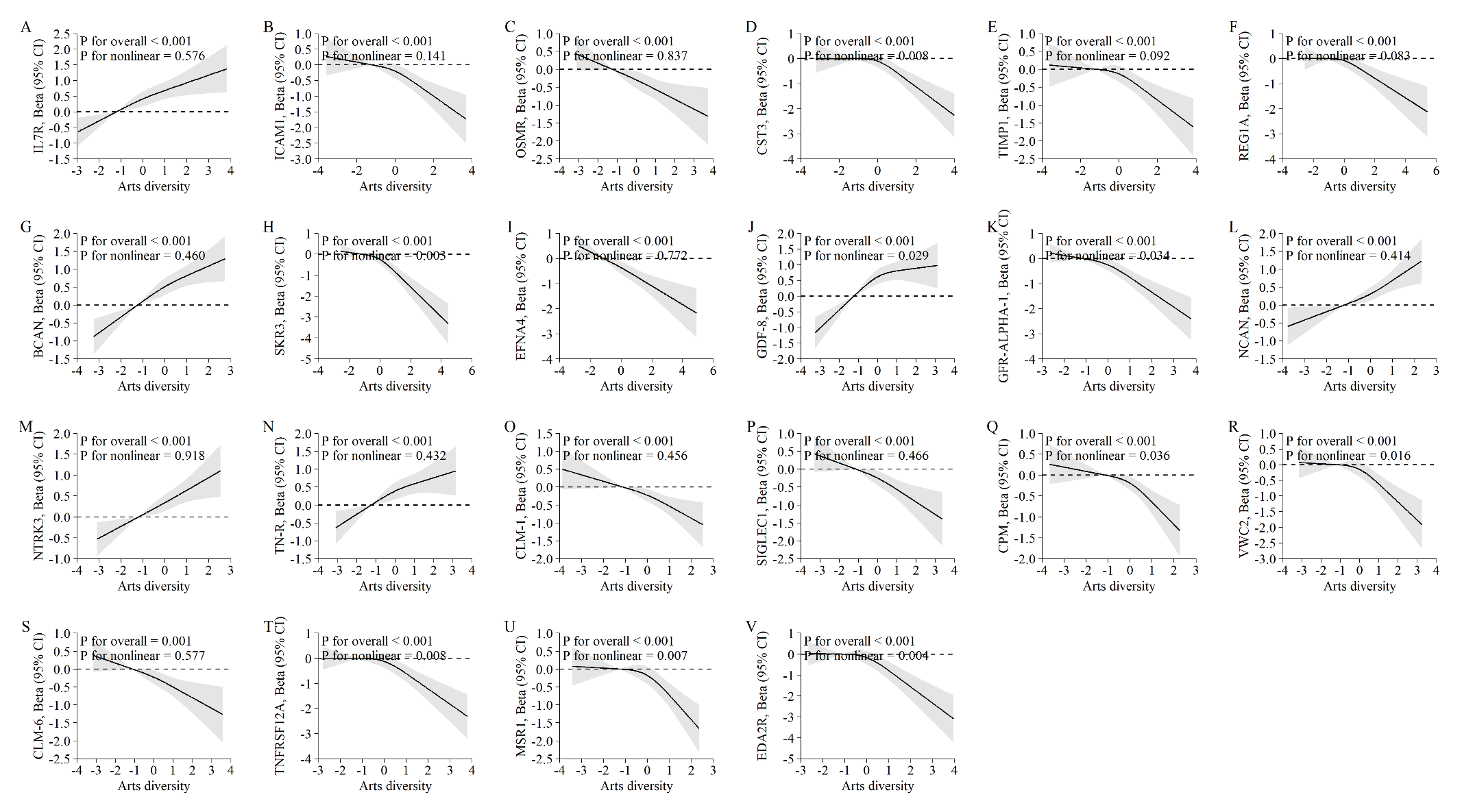

**Figure S4.** Dose-response relationship between arts-index 2 and proteins that were significant after Bonferroni correction.

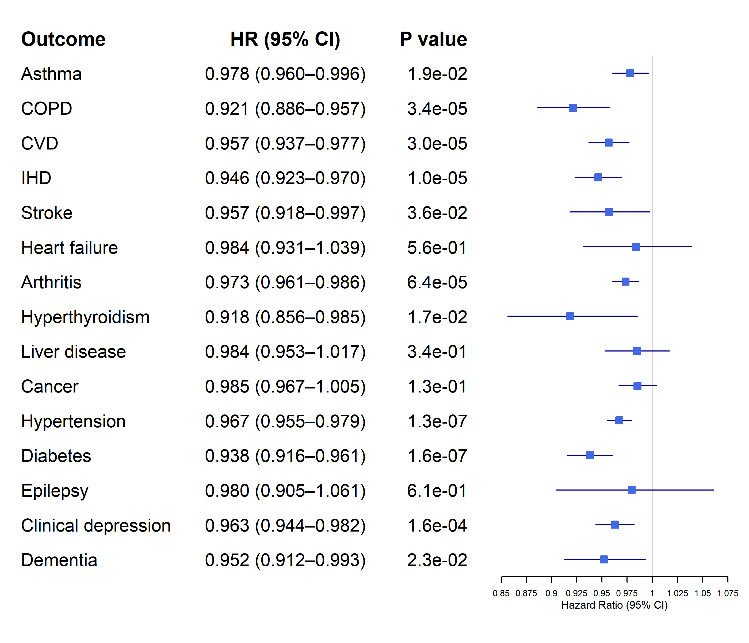

(b)

(c)

(a)

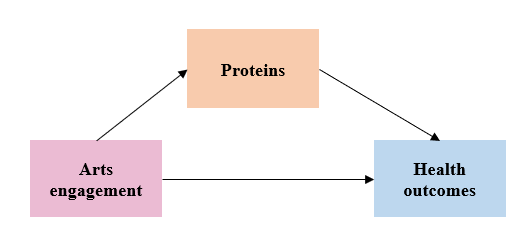

**Multiple mediation analysis**

Arts index-2

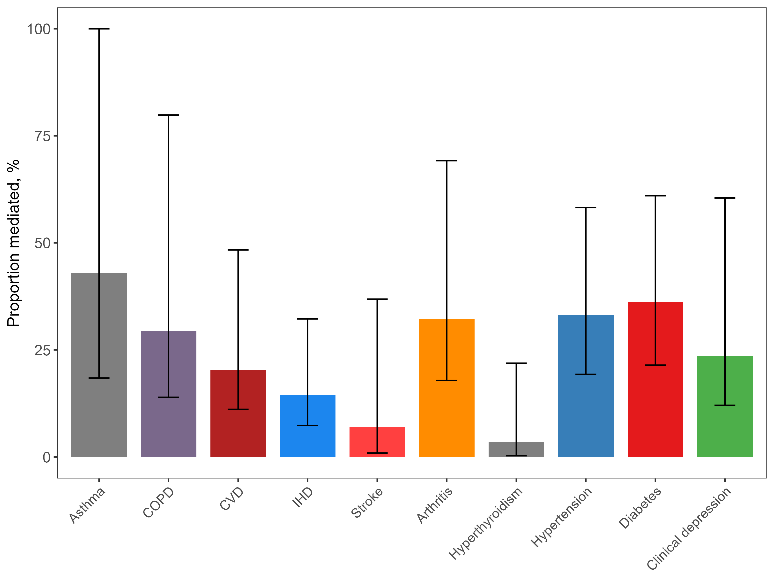

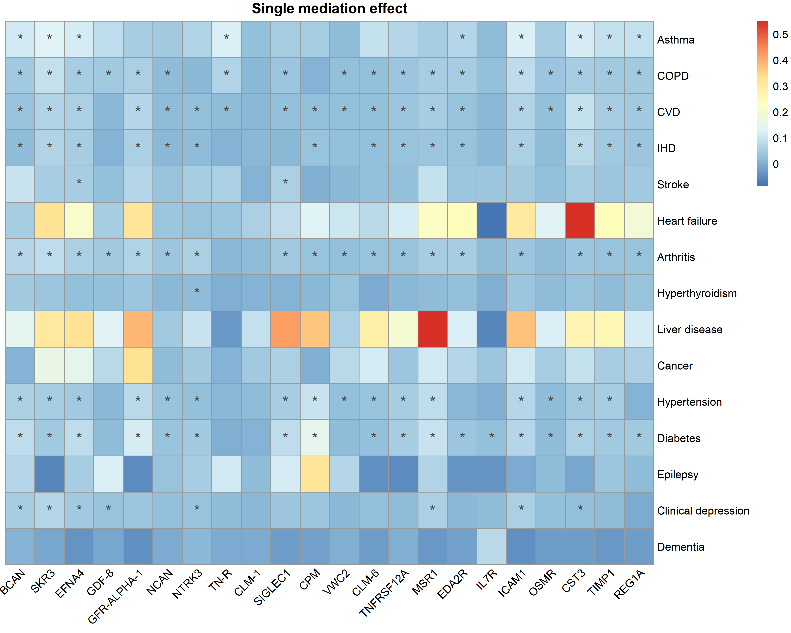

**Figure S5.** Arts index-2 and health outcomes and the role of serum proteins. **(a) Forest plots showing arts index-2 and associations with 15 long-term health outcomes.** Cox regression models were conducted to generate the Hazard ratios and 95% CIs of arts index-2 and health outcomes (except for dementia which used logistic regression). The models were all adjusted for age, age^2^, sex, ethnicity, marital status, education, income, employment status, and area deprivation.
**(b) Single mediation analyses of proteins in the associations between arts index-2 and health outcomes.** The heatmap only shows the percentage of mediation for the health outcomes that had a significant indirect effect of the protein in the associations between arts index-2 and health outcomes. Those proteins that were non-significant across outcomes are not shown. The value is the percentage of mediation of each protein and those which were significant in the mediation analysis are marked as “*”. The mediation models adjusted for age, age^2^, sex, ethnicity, marital status, education, income, employment status, and area deprivation.
**(c) Multiple mediation analyses of all identified significant proteins in the associations between arts index-2 and health outcomes.** The models were all adjusted for age, age^2^, sex, ethnicity, marital status, education, income, employment status, and area deprivation.

**Arts index-2**

**Arts index**

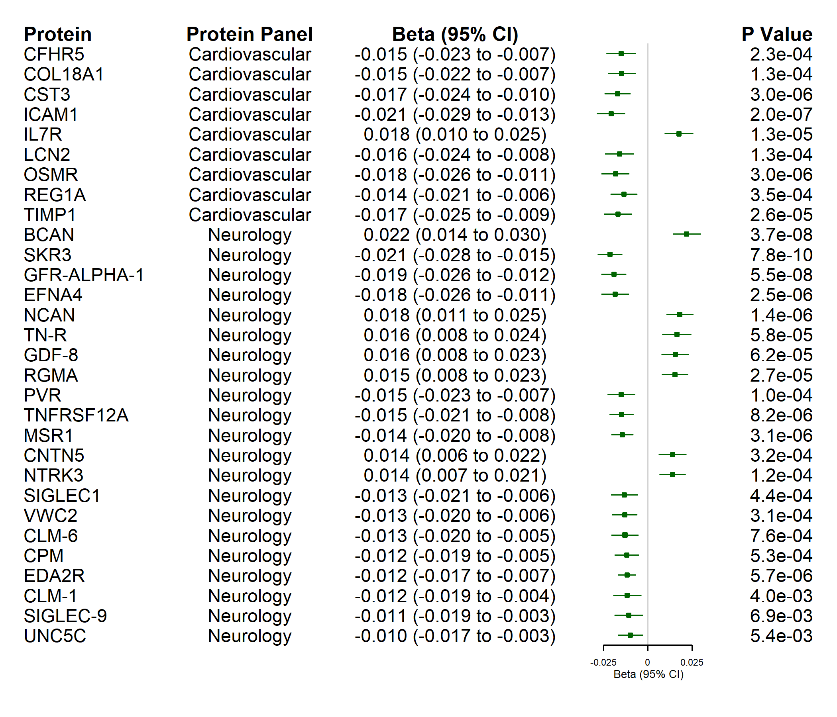

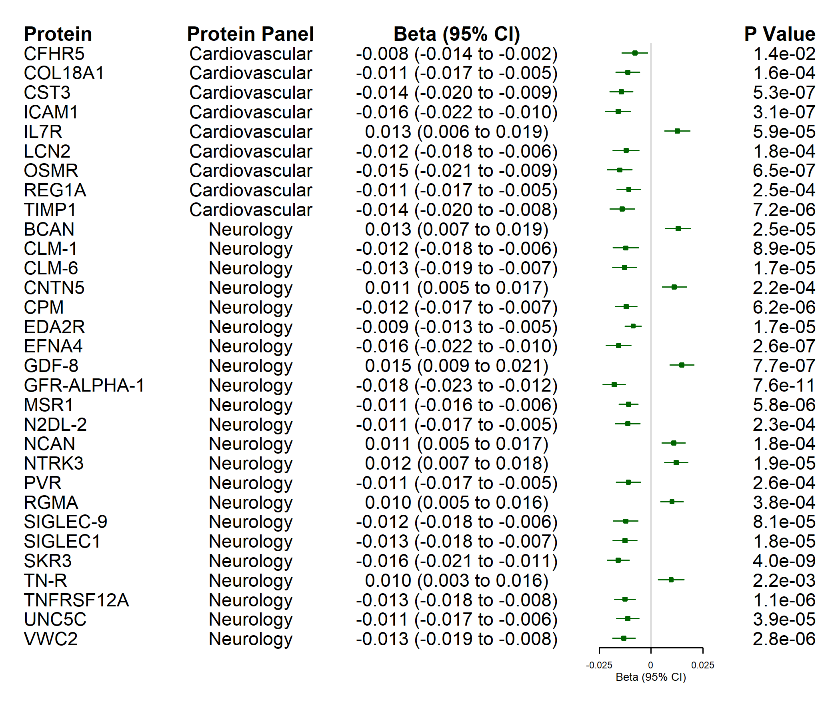

**Figure S6.** Sensitivity analysis for arts index and arts index-2 and serum proteins when excluding those aged 40 years or less.
This figure only provides the associations for the proteins (number of proteins = 30) that were significant after Bonferroni correction for arts index (number of proteins = 21) and arts index-2 (number of proteins = 28). The sample size was 4494 for the cardiometabolic proteins panel and 4565 for neurology proteins panel.
